# Androgen receptor polyQ alleles and COVID-19 severity in men: a replication study

**DOI:** 10.1101/2022.03.25.22271678

**Authors:** Rosario López-Rodríguez, Javier Ruiz-Hornillos, Marta Cortón, Berta Almoguera, Pablo Minguez, María Elena Pérez-Tomás, María Barreda-Sánchez, Esther Mancebo, Lorena Ondo, Andrea Martinez-Ramas, Lidia Fernández-Caballero, the STOP_Coronavirus Study Group, Estela Paz-Artal, Encarna Guillen-Navarro, Carmen Ayuso

**Affiliations:** Department of Genetics & Genomics, Instituto de Investigación Sanitaria-Fundación Jiménez Díaz University Hospital, Universidad Autónoma de Madrid (IIS-FJD, UAM), Madrid, Spain; Center for Biomedical Network Research on Rare Diseases (CIBERER), Instituto de Salud Carlos III, 28029, Madrid, Spain; Allergy Unit, Hospital Infanta Elena, Valdemoro, Madrid, Spain; Instituto de Investigación Sanitaria-Fundación Jiménez Díaz University Hospital, Universidad Autónoma de Madrid (IIS-FJD, UAM), Madrid, Spain; Faculty of Medicine, Universidad Francisco de Vitoria, Madrid, Spain; Medical Genetics Section, Pediatric Department, Hospital Clínico Universitario Virgen de la Arrixaca, Instituto Murciano de Investigación Biosanitaria Virgen de la Arrixaca, Universidad de Murcia (IMIB-Arrixaca, UMU), Murcia, Spain; Health Sciences Faculty. Universidad Católica San Antonio de Murcia (UCAM), Murcia, Spain; Department of Immunology, Hospital Universitario 12 de Octubre, Madrid, Spain; Instituto de Investigación Sanitaria Hospital 12 de Octubre (imas12), Madrid, Spain; Department of Immunology, Ophthalmology and ENT, Universidad Complutense de Madrid, Madrid, Spain

## Abstract

Ample evidence indicates a sex-related difference in severity of COVID-19, with less favorable outcomes observed in men. Genetic factors have been proposed as candidates to explain this difference. The polyQ polymorphism in the androgen receptor gene has been recently described as a genetic biomarker of COVID-19 severity. In this study, we analyzed this association in a large cohort of 1136 men classified into three groups according to their degree of COVID-19 severity, finding a similar distribution of polyQ alleles among severity groups. Therefore, our results do not support the role of this polymorphism as a biomarker of COVID-19 severity.

## Introduction

Since the first outbreak of the COVID-19 pandemic, over 307 million cases of COVID-19 and more than 5.4 million deaths have been confirmed worldwide (https://coronavirus.jhu.edu, *last accessed* in January 2022). A wide body of evidence suggests a sex-related difference in the severity of the disease, with less favorable outcomes observed in men. However, the infection rate, lifestyle factors, age, and comorbidities cannot completely explain these sex-based differences in outcomes (1).

Genetic background has been proposed as an explanatory factor in some of these sex-based differences. Interestingly, various X-linked genes are implicated in the immune response (2), such as loss-of-function *TLR7* variants, which are related to fatal outcomes in young males with COVID-19, or *ACE2*, which is known to mediate the entry of SARS-CoV-2 (2,3). Additionally, the association between the polyQ polymorphism within exon 1 in the androgen receptor (*AR*, Xq12) gene and COVID-19 severity has been recently described (4). This genetic variant consists of a polymorphic CAG repeat segment located at the N-terminal transactivation domain, ranging from 9 to 36 repeats in non-pathological conditions, and its length is inversely correlated with AR activity (5,6). A number of ≥23 CAG repeats was significantly associated with a more severe COVID-19 outcome in an initial cohort of 638 Italian male and female patients; these results were replicated in an independent cohort of 158 Spanish male patients under 60 years of age (4).

Response to androgen hormones (testosterone, androsterone, and androstenedione) is mediated by the AR. Upon androgen binding to the AR, the AR translocates to the nucleus, where it regulates the expression of the androgen-responsive genes (7). Previous studies have suggested a key role of androgens in COVID-19 severity. Lower testosterone concentrations have been related to higher severity and mortality in males with COVID-19 (8,9). Also, the expression of TMPRSS2, a protease that primes the SARS-CoV-2 spike protein, is regulated by elements of the androgen response (10). Interestingly, AR transactivational activity is regulated by the polyQ polymorphism.

In the present study we aimed to test the association between the AR polyQ polymorphism with severity, reported previously (4), in a large cohort of 1136 COVID-19 male patients, stratified by the disease outcome and adjusting by known risk factors such as age, comorbidities and ethnicity.

## Subjects and Methods

### Subjects

This study included 1136 male patients from the STOP_Coronavirus cohort who were infected with SARS-CoV-2 as confirmed by positive PCR. Patients were retrospectively and prospectively enrolled from March to November 2020 and followed-up until February 2021. Patients were recruited from four hospitals in Spain, three of which are in Madrid (Hospital Universitario Fundación Jiménez Díaz [HUFJD], Hospital Universitario Infanta Elena [HUIE], and Hospital Universitario 12 de Octubre [H12O]) and one in Murcia (Hospital Clínico Universitario Virgen de la Arrixaca [HUVA]).

This study was approved by the research ethics committees of each center (Clinical Research Ethics Committee Hospital Universitario Fundación Jiménez Díaz, Fundación Jiménez Díaz Biobank, ref. PIC087-20; Clinical Research Ethics Committee Hospital Universitario Virgen de la Arrixaca, Instituto Murciano de Investigación Biosanitaria Virgen de la Arrixaca Biobank, ref. COVID-19 RMu; and Clinical Research Ethics Committee Hospital 12 de Octubre). The STOP_Coronavirus cohort received Clinical Research Ethics Committee approval PIC087-20 from Hospital Universitario Fundación Jiménez Díaz. Wherever possible, patients provided written or verbal informed consent to participate. Due to the health emergency, the research ethics committees of each center waived the requirement for informed consent for the STOP_Coronavirus cohort. All samples were de-identified (pseudonymized) and clinical data were managed in accordance with national legislation and institutional requirements.Clinical data

Clinical data obtained in HUFJD and HUIE were extracted from individual electronic medical records using big data/artificial intelligence and then reviewed and refined by four independent researchers. At H12O and HUVA, clinical data were manually collected from electronic medical records. Clinical information included primary demographic data, comorbidities, COVID-19 symptoms, laboratory findings, treatments, related complications from COVID-19, ICU admission, and outcome.

Severity stratification was performed using the following criteria: 1) oligosymptomatic, asymptomatic subjects or patients with mild symptoms (fever, cough, sore throat, fatigue, and shortness of breath) who did not require hospitalization (n=176); 2) hospitalized patients not requiring invasive or non-invasive mechanical ventilation (n=718); 3) severe patients admitted to the ICU or similar units (n=242) and requiring intubation, non-invasive ventilation (CPAP/BiPAP), or high-flow nasal cannula (≥16L/min) with clinical signs of severe SARS-CoV-2 infection, including survivors and deceased patients.

### Genotyping of the androgen receptor polyQ polymorphism

Genomic DNA was isolated from EDTA-collected peripheral blood samples using an automated DNA extractor (BioRobot EZ1, QIAGEN GmbH). Genotyping was performed as previously described (11) with minor modifications. The number of CAG repeats (polyQ polymorphism) at the androgen receptor was obtained by PCR using a forward fluorescent primer followed by electrophoresis (ABI3130 sequence, Applied Biosystems). Allele size was calculated using the Genescan Analysis software (Applied Biosystems) and patients were classified as either short (<23 repeats) or long (≥23 repeats) allele carriers.

### Ancestry inference

Principal component analysis (PCA) based on the variance-standardized relationship matrix was used to infer the ancestry of each patient and classify them as one of the selected ancestry groups (European, African, admixed American, and East Asian) using a set of 1000 genome samples (phase 3) as a reference population. For PCA, we used previously collected genetic data from our cohort (unpublished) obtained with Applied Biosystems™ Axiom™ Spain Biobank Array (COL32017 1217, Thermo Fisher Scientific Inc.), which contains758,740 variants. PCA was performed using Plink software, version 1.9 (12). Inferred patient ancestry was used to stratify our population into three groups: Europeans (n=813), admixed Americans (n=194), and others (n=129).

### Statistical analysis

The statistical association of demographic and clinical variables with COVID-19 severity was calculated by Chi-squared (Chi2) test and non-parametric Kruskal-Wallis test for categorical (ethnicity and comorbidities) or continuous variables (age and CAG repeats), respectively.

The association between polyQ alleles (short < 23 or long ≥ 23) and COVID-19 severity was assessed by Chi2 test in 1) the overall COVID-19 cohort (n=1136); 2) patients under 60 years of age (n=522); 3) patients of European ethnicity (n=813); and 4) European patients under age 60 years (n=300).

Multivariate logistic regression analysis was performed to test the association of different demographic (age, facility, ethnicity), clinical (comorbidities), and genetic (polyQ polymorphism) variables with COVID-19 severity. Patients displaying either of the two extreme severity phenotypes were selected for logistic regression analysis, which compared severe and oligosymptomatic patients. Two-tailed p-values below 0.05 were considered statistically significant.

## Results

A total of 1136 male patients with confirmed SARS-CoV-2 infection as indicated by a positive PCR assay were selected from the Spanish STOP_Coronavirus cohort, which comprises more than 3,500 COVID-19 patients. Patients included in the present study were recruited mainly during the first wave of the pandemic in Spain (March to April 2020) from three hospitals located in Madrid (n=991, 87%) and one in Murcia (n=145, 13%).

The average patient age was 60.56 ± 15.37 years, and patients were mostly of European origin (n=813, 71%). Hypertension (41%) and obesity (20%) were the most frequent comorbidities (Table 1). Patients were classified into three severity groups as follows: 1) oligosymptomatic (15.5%); 2) hospitalized patients not requiring substantial respiratory support (63.2%); and 3) severe patients (21.3%).

**Table 1.** Demographic and clinical data among the COVID-19 severity categories for the patients included in the study.

|  | All patients | Oligosymptomatic<sup>1</sup> | Hospitalized<sup>2</sup> | Severe<sup>3</sup> |
| --- | --- | --- | --- | --- |
| Characteristics | n=1136 | n=176 | n=718 | n=242 |
| Age (mean ± SD) * | 60.56 ± 15.37 | 55.25 ± 14.84 | 61.92 ± 15.86 | 60.40 ± 13.37 |
| Ethnicity (EUR/AMR/Others) * | 813/194/129 | 146/20/10 | 511/113/94 | 156/61/25 |
| PolyQ CAG repeats (mean) | 22 | 22 | 22 | 22 |
| Hypertension (n, %) * | 463, 41% | 56, 32% | 305, 42% | 102, 42% |
| Type 2 diabetes (n, %) | 110, 10% | 10, 6% | 73, 10% | 27, 11% |
| Obesity (n, %) * | 231, 20% | 24, 14% | 142, 20% | 65, 27% |
| Cardiovascular disease (n, %) | 139, 12% | 15, 8% | 92, 13% | 32, 13% |
| Neurological disease (n, %) | 50, 4% | 6, 3% | 32, 4% | 12, 5% |
| Respiratory disease (n,%) * | 95, 8% | 6, 3% | 65, 9% | 24, 10% |

<sup>1</sup>Patients showing mild COVID-19 symptoms not requiring hospitalization, seven patients received supplemental oxygen. <sup>2</sup>Hospitalized patients that did not require respiratory support or only received supplemental oxygen. <sup>3</sup>Patients admitted to the ICU and requiring respiratory support by means of intubation, non-invasive ventilation (CPAP/BiPAP), or high flow nasal cannula ( $\geq 16$ L/min). \*Statistical differences among the severity categories ( $p < 0.05$ ) calculated by Kruskal-Wallis test for continuous variables (age and CAG repeats) and $\chi^2$ test for categorical variables (ethnicity and comorbidities). SD, standard deviation; n, number of patients; %, percentage of patients; ICU, intensive care unit; EUR, European ancestry; AMR, Admixed American ancestry.

The mean number of polyQ CAG repeats was 22 (±3) (Table 1), a similar finding to various Mediterranean populations, including one from Spain (13). Allele distribution, coded as shorter (<23 repeats) or longer (≥23 repeats) alleles, did not show significant differences between severity classes in our cohort (Table 2, Chi2 test p>0.05). Moreover, the distribution of longer polyQ alleles was similar among milder cases (oligosymptomatic group, 43%) and more severe patients (39%). The association between longer allele frequency and outcome in patients under 60 years of age was also analyzed (Table 2), as age is a well-known risk factor for severe disease. Similar results were observed for the cohort of patients under 60 years of age, in which 38% to 44% were found to have the longer allele (not significant). As the length of the polyQ CAG polymorphism varies among populations, we analyzed the association of this genetic variant with severity in those patients of European ancestry (n=813 and n=300 for the Europeans under 60 years old), finding similar results to those obtained for the overall cohort (Table 2, p>0.05). In addition, polyQ polymorphism distribution was studied in the two groups with extreme COVID-19 severity, finding a similar distribution of this genetic variant among mild and severe cases in the overall population, among Europeans, and in patients under 60 years of age (Table 2).

**Table 2.** PolyQ alleles of the androgen receptor in the STOP_Coronavirus cohort: distribution among severity categories in male patients.

| Severity | Males |  | p-value | Males <60 yo |  | p-value | EUR males |  | p-value | EUR <60 yo |  | p-value |
| --- | --- | --- | --- | --- | --- | --- | --- | --- | --- | --- | --- | --- |
|  | (n=1136) |  |  | (n=522) |  |  | (n=813) |  |  | (n=300) |  |  |
|  | <23 | ≥23 |  | <23 | ≥23 |  | <23 | ≥23 |  | <23 | ≥23 |  |
| Oligosymptomatic<sup>1</sup> | 100 | 76 |  | 62 | 45 |  | 86 | 60 |  | 51 | 33 |  |
|  | (57%) | (43%) |  | (58%) | (42%) |  | (59%) | (41%) |  | (61%) | (39%) |  |
| Hospitalized<sup>2</sup> | 412 | 306 | 0.55<sup>a</sup> | 173 | 137 | 0.55<sup>a</sup> | 301 | 210 | 0.75<sup>a</sup> | 95 | 71 | 0.85<sup>a</sup> |
|  | (57%) | (43%) | 0.37<sup>b</sup> | (56%) | (44%) | 0.56<sup>b</sup> | (59%) | (41%) | 0.56<sup>b</sup> | (57%) | (43%) | 0.93<sup>b</sup> |
| Severe<sup>3</sup> | 148 | 94 |  | 65 | 40 |  | 97 | 59 |  | 30 | 20 |  |
|  | (61%) | (39%) |  | (62%) | (38%) |  | (62%) | (38%) |  | (59%) | (41%) |  |

<sup>1</sup>Patients showing mild COVID-19 symptoms not requiring hospitalization, seven patients received supplemental oxygen. <sup>2</sup>Hospitalized patients that did not require respiratory support or only received supplemental oxygen. <sup>3</sup>Patients admitted to the ICU requiring respiratory support by intubation, non-invasive ventilation (CPAP/BiPAP), or high flow nasal cannula (≥16 L/min). ICU, intensive care unit; EUR, European ancestry; yo, years old. <sup>a</sup>Chi<sup>2</sup> test was not significant (p≥0.05) for the allelic distribution between the three severity groups. <sup>b</sup>Chi<sup>2</sup> test was not significant (p≥0.05) for the allelic distribution between oligosymptomatic and severe patients (2X2 table).

The association of the polyQ alleles with severity grade in the two extreme groups (severe vs. oligosymptomatic) was also assessed in a multivariate logistic regression analysis in which demographic and clinical variables, including comorbidities, were also considered (Table 3). The results indicated that enrollment centers, ethnicity, age, and obesity were significantly associated with severe COVID-19 in our cohort. However, longer polyQ alleles were not associated with severe COVID-19 (p≥0.05) after adjusting for different covariables (Table 3). Similar results were obtained when the two groups with extreme severity were limited to patients of European ancestry (Table 3).

**Table 3.** Multivariate logistic regression analysis: demographic and clinical variables associated with severe illness.

| Variable | Total<sup>a</sup> |  | Europeans<sup>b</sup> |  |
| --- | --- | --- | --- | --- |
|  | OR (95% CI) | p-value | OR (95% CI) | p-value |
| Center |  | <0.001 |  | 0.40 |
| HUFJD | 0.10 (0.02 – 0.31) | <0.001 | - | - |
| HUIE | 0.10 (0.02 – 0.31) | <0.001 | - | - |
| HUVA | 0.25 (0.05– 1.08) | 0.06 | - | - |
| Ethnicity |  | <0.001 | - | - |
| EUR | 0.39 (0.14 - 1.10) | 0.07 | - | - |
| AMR | 1.73 (0.57 - 5.26) | 0.33 | - | - |
| Age (years) | 1.05 (1.02 - 1.07) | <0.001 | 1.05 (1.02 - 1.07) | <0.001 |
| PolyQ long allele ($\geq 23$) | 0.66 (0.40 - 1.07) | 0.10 | 0.70 (0.39 - 1.25) | 0.23 |
| Hypertension | 0.97 (0.54 - 1.72) | 0.91 | 1.04 (0.54 - 1.99) | 0.91 |
| Type 2 diabetes | 0.88 (0.32 - 2.37) | 0.79 | 1.01 (0.33 – 3.11) | 0.98 |
| Obesity | 2.97 (1.60 - 5.50) | <0.001 | 2.97 (1.51 - 5.84) | 0.002 |
| Cardiovascular disease | 0.79 (0.33 - 1.88) | 0.59 | 0.82 (0.32 – 2.10) | 0.69 |
| Neurological disease | 0.43 (0.09- 2.05) | 0.29 | 0.44 (0.09- 2.11) | 0.31 |
| Respiratory disease | 1.89 (0.55 - 6.49) | 0.31 | 1.60 (0.44 – 5.88) | 0.48 |

<sup>a</sup>A total of 418 patients were included, 352 without missing value in one or more of the variables (182 severe and 170 oligosymptomatic patients). <sup>b</sup>A total of 302 patients were included, 251 without missing value in one or more of the variables (110 severe and 141 oligosymptomatic patients). EUR, European ancestry; AMR, Admixed American ancestry.

## Discussion

Several studies have reported higher percentages of hospitalization, morbidity, and mortality in males with COVID-19 compared to females. Different biological mechanisms such as the immune response, viral entry, or sex hormones have been proposed to explain the sex bias in COVID-19 severity and mortality.

In this study, we analyzed the value of the polyQ polymorphism of the AR, which is located on chromosome Xq12, as a biomarker of severity in a cohort of 1136 male patients with COVID-19. Our analysis only includes males so as to prevent the confounding effect of X chromosome inactivation of this locus in females. An important number of immune-related genes and regulatory elements involved in both the innate and adaptive immune response are encoded on the X chromosome, and the AR gene is located at Xq12. Inactivation of one of the two X chromosomes makes females functional mosaics for most of the genes located on X chromosome. Strikingly, some of these immune genes, such as TLR7 or TLR8, escape inactivation (14,15). Such phenomena could confer an immunological advantage in females’ response to infectious diseases. Loss-of-function variants in TLR7 have been associated with fatal COVID-19 outcomes in a small case series of four male patients (3). However, the occurrence of deleterious rare variants in genes at the X chromosome may explain only a small percentage of the higher risk observed in males.

A growing body of evidence suggests that androgens affect COVID-19 severity (1,2,16). Lower testosterone concentrations have been related to higher severity and mortality in males with COVID-19 (8,9). Androgens increase the expression of TPMRSS2 (17,18) and are thought to enhance the activity of ACE2 (19), thus facilitating the entry of SARS-CoV-2. Additional data suggest a role played by androgens in COVID-19 outcome, as the level of testosterone is lower in males with a more severe course compared to those with milder illness (8,9). Therefore, androgen sensitivity may be a critical factor in COVID-19 disease severity. AR is the key regulator in the response to androgens as it modulates the expression of the androgen-responsive genes (7). Of note, AR gene expression correlated inversely with the length of the polyQ polymorphism (5). Based on this evidence, the polyQ polymorphism represents an attractive biomarker of severity in COVID-19. However, our results do not support the findings of *Baldassarri et al* who found more severe outcomes in patients with ≥23 CAG repeats (4).

In the present study, most patients were recruited during the first wave of the pandemic (March to April 2020) from hospitals located in Madrid (n=991). During this period, health-care systems were overwhelmed worldwide, and clinical criteria for hospitalization have varied over the course of the pandemic. Therefore, we hypothesize that our control group (oligosymptomatic) may include patients with more mild-to-moderate symptoms compared to those selected by *Baldassarri et al* (4). However, there is no information in the previous study on enrollment dates and, similarly to the present study, the single inclusion criterion for the control group was the absence of required hospitalization. Therefore, even though the present study includes a larger cohort of patients, we cannot rule out the possibility that the differences observed in both studies are due to discrepancies between control groups.

Furthermore, a high rate of false positives was seen during the first wave of the pandemic due to the use of serological tests to detect SARS-CoV-2 infection. Therefore, statistical analysis in our cohort was performed in patients with a positive PCR test to avoid false positive results, particularly in the control group. Of note, the mean number of polyQ CAG repeats was the same among severity categories in our cohort. Similar to the discovery study (4), we have performed a multivariate logistic regression analysis to reduce the effect of confounding factors (comorbidities) on the association between polyQ allele and COVID-19 severity. After this analysis, we were not able to replicate the effect of the polyQ polymorphism on COVID-19 disease in our larger cohort of patients. However, some aspects in the design of our study differ from the study performed previously (4), mainly related to the clinical characterization of the pre-existing conditions of both cohorts. Therefore, these differences may account to the differential outcome obtained regarding the value of the polyQ polymorphism as a COVID-19 severity biomarker.

In conclusion, our results do not support the value of the polyQ polymorphism in the AR gene as a severity biomarker of COVID-19. Additional studies are needed to clarify the mechanism underlying the association between androgens and COVID-19 severity.

## Data Availability

All data produced in the present study are available upon reasonable request to the authors

## Data Availability Statement

The data supporting the findings of this study are available from the corresponding author on reasonable request.

## Acknowledgements

We thank patients for their generous contribution. We thank the collaboration of FJD-Biobank, registered on the Registro Nacional de Biobancos (B.0000647) supported by the ISCIII (proyecto PT20/00141) and BIOBANC-MUR, registered on the Registro Nacional de Biobancos (B.0000859) supported by the ISCIII (proyecto PT20/00109), IMIB-Arrixaca and Consejería de Salud de la CARM. We are also grateful to Almudena Avila-Fernandez and María José Trujillo-Tiebas for thoughtful discussions. We recognize Ascensión Gimenez for her technical assistance. Lastly, we thank Oliver Shaw for editorial assistance.

## The STOP_Coronavirus Study Group

- **Associated Clinical and Researchgroup of University Hospital Fundación Jiménez Díaz**: Miguel Górgolas, Alfonso Cabello, Germán Peces Barba, Sara Heili, César Calvo, M^a^ Dolores Martín Ríos, Arnoldo Santos, Olga Sánchez-Pernaute, Lucía Llanos, Sandra Zazo, Federico Rojo, Felipe Villar, Raimundo de Andrés, Ignacio Jiménez Alfaro, Ignacio Gadea, Celia Perales, Ruth Fernández Sanchez, Laura Marzal Gordo, Cristina Villaverde, Inés García Vara, Yolanda Cañadas Juarez. Ignacio Mahillo, Antonio Herrero, Juan Carlos Taracido.
- **Associated clinical group of University Hospital Infanta Elena**: Ángel Jiménez, María Herrera Abián, Mercedes García Salmones, Lidia Gagliardi Alarcon, María Rubio Oliveira, CarlosFabian Castaño Romero, Carlos Aranda Cosgaya, Virginia Víctor Palomares, Leticia García Rodríguez, MariaSanchez Carpintero Abad, M^a^ Carmen García Torrejón.
- **Associated Clinical and Research group of University Hospital 12 de Octubre**: Alberto Utrero-Rico, Mario Fernández-Ruiz, Octavio Carretero, José María Aguado, Rocio Laguna-Goya.
- **Associated Clinical and Researchgroup of IMIB-Arrixaca /University Hospital Virgen de la Arrixaca**: Elisa García-Vázquez, Rubén Jara-Rubio, José A Pons-Miñano, Juana M. Marín-Martínez, M. Teresa Herranz-Marín, Enrique Bernal-Morell, Josefina García-García, Juan de Dios González-Caballero, M. Dolores Chirlaque-López, Alfredo Minguela-Puras, Manuel Muro-Amador, Antonio Moreno-Docón, Genoveva Yagüe-Guirao, José M. Abellán-Perpiñán, Jorge E. Martínez-Pérez, Fernando I. Sánchez-Martínez.

## Authors’ contributions

C. A. conception of the study. R. L-R., J. R.-H., M.E. P.-T., M. B.-S., E. M., E. P.-A., E. G.-N. and C.A. provided samples and clinical data. R. L.-R., J. R.-H., M. C., B. A., P. M. and C.A. reviewed the clinical data and classify patients in severity categories. L. O., A. M.-R. and L. F.-C. performed the genotyping of the samples. R. L.-R. and C. A. wrote the initial manuscript and performed statistical analyses. All authors contributed to the review and approval of the final version of the manuscript.

## Ethical Approval

This study was approved by the research ethics committees of each center (CEIm HUFJD, FJD-Biobank, ref. PIC087-20; CEIm HUVA, IMIB-Arrixaca Biobank, ref. COVID-19 RMu; and CREC H12O). The STOP_Coronavirus cohort received CEIm approval PIC087-20 from HUFJD.

